# Effect of Female Age on Infertility Diagnostic Factors and In-vitro Fertilization Treatment Outcomes: A Single-center Retrospective Cohort Study

**DOI:** 10.1101/2021.01.04.21249246

**Authors:** Zhouxuan Li, Songyuan Tang, Shan Liu, Huan Xu, Zhen Lei, Ying Zhong

## Abstract

**Background:** Infertility has become an important issue in modern world because of the increasing number of infertile couples around the world. Advanced maternal age was considered to be a main factor of infertile problem. With second child policy published in China and more women in China intend to seek help for infertility problem, this study provided information of fertility diagnostic factors and IVF treatment outcomes of female IVF patients in different age groups, which can be a guidance for infertility diagnostic and treatment.

**Methods:** A retrospective cohort study was conducted to IVF patient data collected by Jinjiang District Maternal and Child Health Hospital, Chengdu, China. Clinical and laboratory data of 45,743 fresh, autologous IVF cycles from January 2008 to September 2017 were included in the analysis. The diagnostic factors and treatment outcomes were analyzed for different age groups (age<35, n=30,708; age 35-41, n=11,921 and age≥42, n=3,114) as well as further stratified advanced age groups (age 42, n=933; age 43, n=744; age 44, n=556; age≥45, n=881). The characteristics including number of previous cycles, duration of infertility, BMI, basic FSH, basic AFC, AMH, retrieved oocyte number, fertilized oocyte number, transferred embryo number, baby number and economic cost were stratified according to patient age.

**Results:** The basic characteristics of obesity rate, basic FSH and cancellation rate of IVF cycles in ≥42 years old group were significantly higher than other groups (p<0.01). Basic AFC, AMH, retrieved oocyte number, fertilized oocyte number and transferred embryo number in ≥42 years old group were significantly lower than other groups (p<0.01). Diagnostics characteristics and IVF-ET outcome declined significantly when maternal age increased (p<0.05). In the meanwhile, a preliminary analysis of cost per cycle showed that cost per child increased with patient’s age increase.

**Conclusion:** Although with increasing number of advanced maternal age IVF cycles, the age group of ≥42 years would intend to get unsatisfied outcome and higher cost per child. More guidance and considerations should be focused on encouraging earlier age treatment of infertility.

**Plain English summary:** With more and more women in the global range choose to get late pregnancy because of changes in society and economy, age has become an unavoidable factor in infertility diagnostic and treatment. Advanced age women may experience more infertility problems and negative IVF outcomes. A better understanding of the effect of maternal age on infertility would offer help to both diagnostic and treatment of IVF patient. This study conducted a single-center retrospective cohort analysis to female patients of different age groups and found that women with more advanced age (≥42) would be more easily to experience unsatisfied IVF outcome and higher economic cost to obtain one child. After the publication of second child policy in China in 2013, the number of advanced age patients increased, the necessity of special guidance for AMA patient may need to be taken into consideration.

## Background

Infertility is a global health problem affecting 15% of couples in the world [1]. It is defined as the “inability to conceive within 2 years of exposure to pregnancy” (not pregnant, sexually active, non-contracepting and non-lactating) among women of reproductive age (15–49) [2]. In China, the prevalence of infertility among women of reproductive age attempting to become pregnant was estimated to be 25.0%, and this percentage is known to increase with age [3]. In vitro fertilization and embryo transfer (IVF-ET) and its related technologies are now popular treatments to different infertile problems [4]. Age of reproduction is considered as a key factor which influences IVF prognosis and treatment outcome [5, 6]. Advanced maternal age (AMA) (age≥35 year) is associated with a decline in both ovarian reserve and oocyte competence [7]. And the declines in oocyte yields and oocyte quality are the primary reasons for deteriorating IVF outcomes with advancing female age [8]. Aneuploidy is another major cause of infertility and pregnancy failure, and the proportion of more complex aneuploidy increasing with age [9, 10].

In global range, more and more families in developed countries tend to postpone childbirth behaviors due to changes of educational, medical, social, economic aspects and so on [11-13]. Women above age 40 years has become the most rapidly growing age group of having child, and the age of IVF-ET populations has increased in accordance [7, 14]. It was reported that the proportion of women aged ≥40 years undergoing nondonor ART (Assisted Reproductive Technology) increased from 23.2% in 2010 to 24.0% in 2011 [15]. In China, the proportion of the advanced maternal age (AMA) (age≥35 year) woman gave first birth were 2.96% and 8.56% in 1996 and 2007 respectively, which illustrates an increase trend of childbirth age [16]. This proportion continued to increase after China rolled out the Two□child Policy (one couple is allowed to have the second child) across the country [17].

Although much research has obtained the conclusion that the maternal age would influence IVF treatment outcome, most of their sample sizes are relatively small and even fewer analysis were conducted to AMA age group or even older age groups. The aim of this study is to reveal the trends of female patient’s characteristics and IVF treatment outcomes changing with age, and provided more specific statistical results in each age group especially in advanced maternal age groups (≥35) and even ≥42 years old up to 47 years old. Together with the preliminary analysis for the economic cost of IVF-ET cycle in different age groups, the results of this study can provide some information and guidance to the consultation and management of IVF-ET treatment for different age group patients.

## Methods

### Study design and setting

The retrospective study was conducted to the clinical and laboratory medical records of 45,743 fresh, autologous IVF cycles (53,384 cycles before data preprocessing) in IVF center of Jinjiang District Maternal and Child Health Hospital in Chengdu, China. Data was collected in the period from January 2008 until September 2017.

### Data inclusive/exclusive criteria

Clinical and laboratory medical records of female patients who accepted IVF treatment in single IVF center were extracted from the hospital dataset. Cycles with fresh embryo transfer and autologous oocytes were included in the study. Observations with extreme or normal values were reviewed and investigated, cycles with mistakes or missing values in main analysis variables were excluded from the study. After data preprocessing, the total number of cycles decreased from 53,384 to 45,743 (age<35, n=30,708; age 35-41, n=11,921 and age≥42, n=3,114).

### Characteristics and outcome variables analyzed

The clinical and laboratory treatment protocols included controlled ovarian stimulation (COS), oocyte retrieval, IVF, and embryo transfer procedures and followed standard protocol descriptions [18, 19].

The characteristics of patient in this study include age, BMI (Body Mass Index), number of unpregnant year, ovarian reserve testing parameters (basic AFC (Antral Follicle Count), AMH (Anti-Müllerian Hormone) and basic FSH (Follicle Stimulating Hormone)), number of oocytes retrieved, number of fertilized oocytes, number of blastocysts and number of embryos transferred.

The outcome variables include biochemical pregnancy, clinical pregnancy, live birth and multiple pregnancy. Positive pregnancy is defined as a serum HCG (Human Chorionic Gonadotropin) concentration of ≥10 IU/l on day 12 after transfer. Clinical pregnancy was defined as the presence of a gestational sac and visualization of fetal heartbeat by ultrasound 2 weeks later. Biochemical pregnancy is defined as *β*-HCG test positive but pregnancy fail to proceed to clinical pregnancy [20]. Live birth is defined as at least one live-born child delivered at >□24 weeks gestation [21]. Multiple pregnancy is defined as number of live birth baby larger than 1.

### Data analysis

Data was preprocessed and analyzed by RStudio version 3.5.0. The differences of characteristics and IVF outcomes among different age groups were tested by chi-square and one-way ANOVA test with multiple comparison. The trend of variables changing with age was tested by Mann-Kendall trend test [22]. P-value <0.05 was considered to be statistically significant.

## Results

Data of 53,384 cycles were collected from January 2008 to September 2017. After investigated the observations with extreme or normal values, 7641 cycles were detected to be with mistakes or with missing value in main variables. 45,743 cycles were left for the following analysis. We first divided data into three age groups, <35 years old, 35-41 years old and ≥42 years old. The descriptive analysis of ovarian reserve testing factors and IVF outcome factors are presented in Table 1. For three age groups, there were 30,708 cycles in <35 years old group, 11,921 cycles in 35-41 years old group and 3,114 cycles in ≥42 years old group. According to the guidance of obesity definition by BMI, obese range would be BMI≥30 [23]. In our research, Basic AFC <5 was considered as low AFC. Since the testing of AMH was started after 2012 in the hospital, smaller number of cycles were included in the analysis of AMH.

**Table 1.**
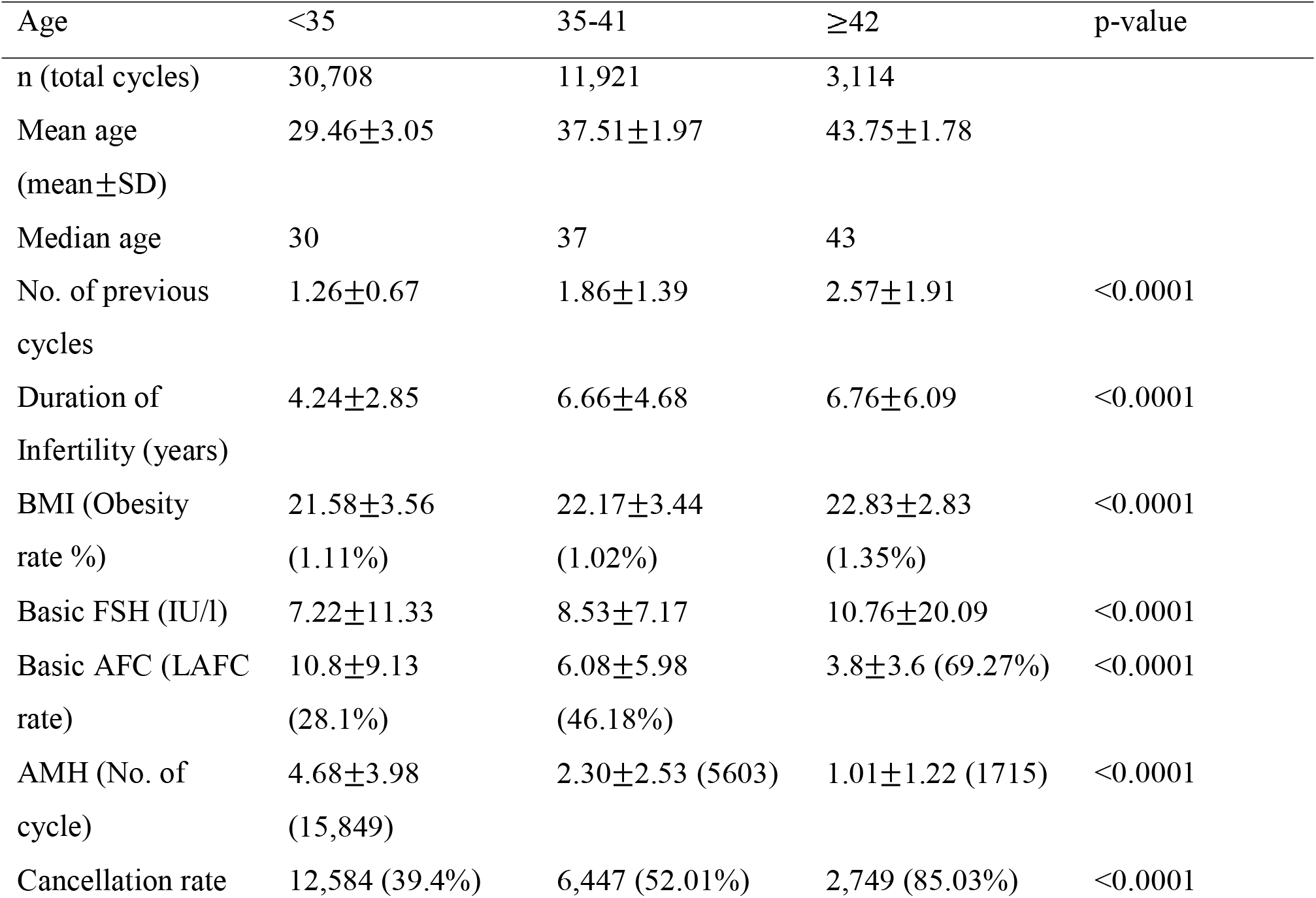
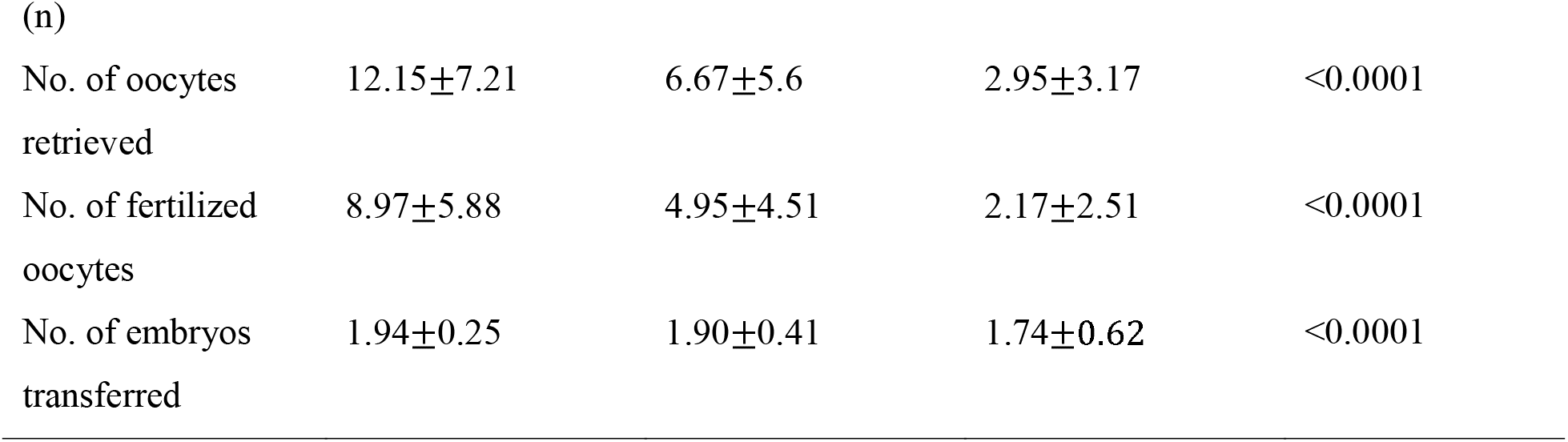
Descriptive analysis of ovarian reserve testing factors and IVF outcome factors in three age groups, <35 years, 35-41 years and ≥42 years.

The number of previous cycles increased with patients’ age increase. And there were significant differences among three groups. For other factors related to IVF outcome, the descriptive analysis also demonstrated that with patients’ age increase, the ovarian reserve testing factors, basic FSH, basic AFC and AMH were getting worse. Specifically, mean basic FSH increased and the mean of basic AFC decreased. Taking the standard of <5 AFC is low AFC, the low AFC rate of three groups increased significantly (from 28.1% to 69.27%). Besides, AMH decreased with age of patient increase. And the three factors were significantly different among the three age groups. As for the IVF outcomes, the cancellation rate of IVF cycle increased (from 39.4% in age group <35 years to 85.03% in age group ≥42 years), number of retrieved oocyte decreased (from 12.15±7.21 in age group <35 years to 2.95±3.17 in age group ≥42 years), number of fertilized oocytes decreased (from 8.97±5.88 in age group <35 years to 2.17±2.51 in age group ≥42 years), and number of transferred embryos decreased (from 1.94±0.25 in age group <35 years to 1.74±0.62 in age group ≥42 years) with age of patient increase. For all the factors, the three age groups were significantly different from each other.

Data of ≥42 years group was further stratified into <42 years, 42 years, 43 years, 44 years and ≥45 years. Table 2 shows the characteristics of different age groups. Mann-Kendall trend test was conducted to test the trend of No. of retrieved oocytes, No. of fertilized oocytes, No. of blastocytes and No. of transferred embryo with relation to age.

**Table 2.** characteristics of research factors of different age groups, <42 years, 42 years, 43 years, 44 years and ≥45 years

| Age | <42 | 42 | 43 | 44 | ≥45 | p-value |
| --- | --- | --- | --- | --- | --- | --- |
| n (total cycles) | 42,629 | 933 | 744 | 556 | 881 |  |
| Basic FSH (IU/l) | 7.59±9.92 | 10.29±7.73 | 11.91±39.29 | 10.14±8.09 | 10.94±8.17 | <0.0001 |
| Basic AFC | 9.58±8.60 | 4.39±4.16 | 3.77±3.49 | 3.95±3.43 | 2.98±2.90 | <0.0001 |
| Cancellation rate (n) | 18,404<br>(43.17%) | 763<br>(81.78%) | 605 (81.32%) | 472<br>(84.89%) | 803<br>(91.15%) | <0.0001 |
| No. of oocytes retrieved* | 10.63±7.24 | 3.62±3.60 | 3.24±3.17 | 2.80±3.06 | 2.02±2.26 | <0.0001 |
| No. of fertilized oocytes* | 10.61±7.22 | 3.62±3.60 | 3.24±3.18 | 2.80±3.07 | 2.03±2.26 | <0.0001 |
| No. of blastocyst* | 7.85±5.83 | 2.68±2.87 | 2.37±2.47 | 2.04±2.46 | 1.49±1.79 | <0.0001 |
| No. of embryos transferred* | 1.93±0.30 | 1.82±0.54 | 1.77±0.61 | 1.70±0.65 | 1.58±0.73 | <0.0001 |

\* means p-value of Mann-Kendall trend test less than 0.0001. Z values are -110.53, -110.36, -100.67 and -42.48 for No. of oocytes retrieved, No. of fertilized oocytes, No. of blastocysts and No. of embryo transferred respectively. Negative z-values demonstrate negative relationship with age.

Basic FSH increase with age from <42 years old to 43 years old, and slightly decreased at 44 years old, and continue with an increasing trend from 44 to 45 years old. Basic AFC decreased with age increase. Cancellation rate of IVF cycle increased with age increase. Number of retrieved oocytes, number of fertilized oocytes, number of blastocyst and number of embryos transferred decreased with patients’ age increase. And all the values in different age groups were significantly different.

Table 3 shows the IVF-ET pregnancy outcomes analysis of different age groups. Embryo transfer cycles were extracted from the data and were divided into 8 age groups including <35 years old, 35-41 years old, 42, 43, 44, 45, 46 and 47 years old.

**Table 3.**
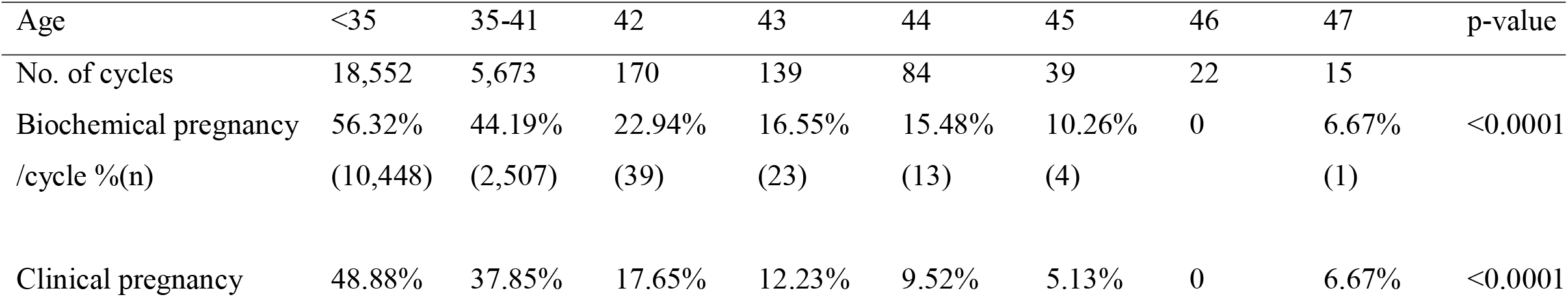
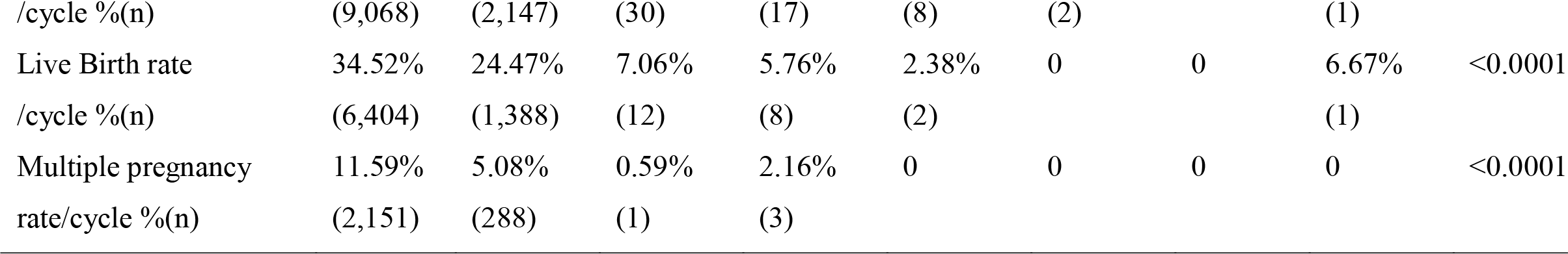
IVF-ET pregnancy outcome of different age groups of <35, 42, 43, 44, 45, 46, 47

Biochemical pregnancy rate was 56.32% in <35 years old group, and decreased with age increase, from 44.19% in age 35-41 years old group to 10.26% in 45 years old group. There was no observation in 47 years old group and 1 observation in 48 years old group. Clinical pregnancy rate of IVF cycles decreased with patients’ age increase. From 48.88% in <35 years old group to 5.13% in 45 years old group. There was no observation in 46 years old group and 1 observation in 47 years old group. Live birth rate decreased with patients’ age increase. From 34.52% in <35 yeas old group to 2.38% in 44 years old group. There was no observation in 45- and 46-years old group and 1 observation in 47 years old group. Multiple pregnancy rate decreased with age increase from <35 years old to 42 years old. However, the observation number decreased tremendously after 41 years old. All the values were significantly different among different age groups.

Table 4 shows the preliminary statistics analysis of economic cost. Draft cost per cycle was estimated for each IVF cycle in different age groups including <35, 42, 43, 44 and ≥45 years old. Although cost per cycle decreases with age increase, cost per child increases largely with maternal age increase (roughly cost per child is 33,514.02 RMB in <35 years old group and 1357,154.07 RMB in ≥45 years old group).

**Table 4.**
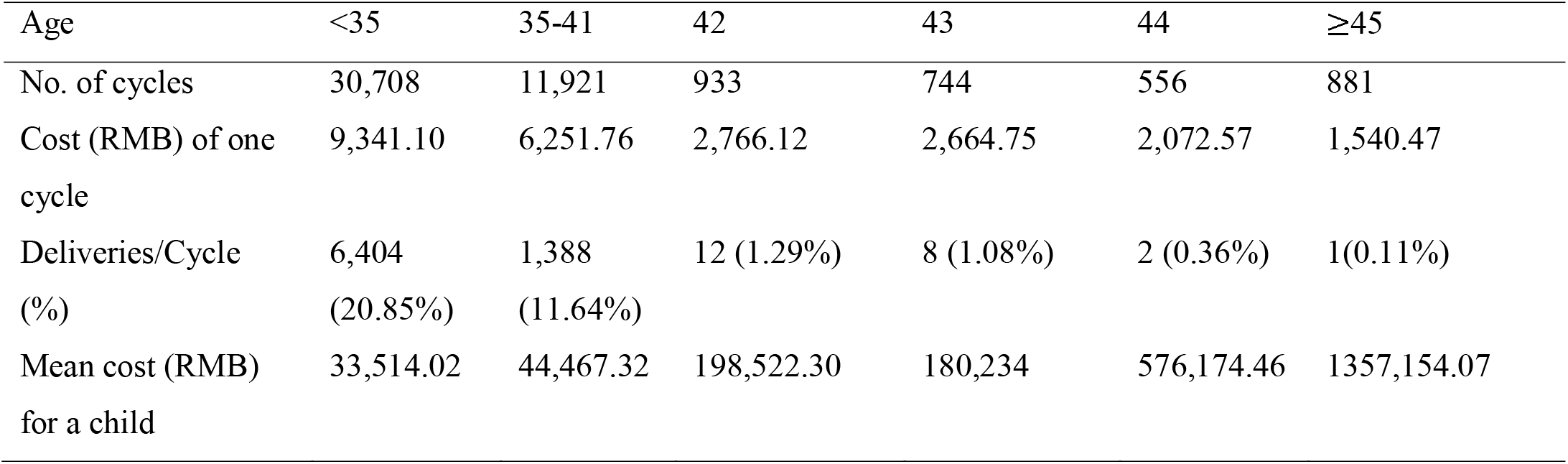
IVF cycle cost for different age groups of <35, 42, 43, 44 and ≥45 years old

| Age | <35 | 35-41 | 42 | 43 | 44 | ≥45 |
| --- | --- | --- | --- | --- | --- | --- |
| No. of cycles | 30,708 | 11,921 | 933 | 744 | 556 | 881 |
| Cost (RMB) of one cycle | 9,341.10 | 6,251.76 | 2,766.12 | 2,664.75 | 2,072.57 | 1,540.47 |
| Deliveries/Cycle (%) | 6,404 (20.85%) | 1,388 (11.64%) | 12 (1.29%) | 8 (1.08%) | 2 (0.36%) | 1(0.11%) |
| Mean cost (RMB) for a child | 33,514.02 | 44,467.32 | 198,522.30 | 180,234 | 576,174.46 | 1357,154.07 |

## Discussion

In this retrospective cohort study, 45,743 fresh, autologous IVF cycles were included. Clinical and laboratory medical records of female patients were analyzed and compared among different age groups. Previous research has shown the declines in oocyte yields and oocyte quality are the primary reasons for deteriorating IVF outcomes with advanced female age [24, 25]. Multinomial regression revealed an association between female age and diagnosis of female infertility [26]. For female IVF patients, follicle stimulating hormone (FSH) and anti-mu□llerian hormone (AMH) are the two most frequently utilized ovarian reserve tests that serve as important markers related to successful IVF outcomes [27]. It is suggested that AMH >0.8-1.0 ng/ml is indicative of normal ovarian reserve, while FSH >10 mIU/ml is indicative of diminished ovarian reserve [28]. Age showed a significant negative correlation with AMH level and AFC. There was a positive correlation between age and FSH [29, 30]. Researchers have found that low level AMH associated with lower live birth rate, on the other hand, low level FSH associated with higher live birth rate. It is also shown that diminished ovarian reserve, which will lead to poor response in IVF, is associated with reduced baseline serum AMH concentration [28, 31, 32]. M. Szamatowicz and D. Grochowski’s research has shown that the clinical pregnancy rate per transfer and the embryo implantation rate declined significantly from 29.4% and 18.9% in women <30 years to 19.8% and 14.3% in patients between 30 and 35 years, 17.1% and 9.0% between 36 and 39 years and to 12.8% and 7.4% in those aged ≥40 years. The spontaneous abortion rate was 14.9%, 16.5%, 22.4% and 33.2%, respectively [33]. ART technology and medical treatment can enhance the chance of pregnancy in young women groups [34]. However, the circumstances are not the same for older women. Loss of ovarian reserve and poor performance with fertility-enhancing medications can contribute to the failure of IVF cycle in AMA women, and even when normal-appearing blastocysts are obtained, trophectoderm biopsies have shown an increasing rate of aneuploidy in the embryos of older women [35]. In the 8-country fertility survey, the research of cost effective of IVF in different age groups shows that the cost effectiveness per live birth of blastocyst transfer is $39,795, $63,191 and $126,930 for age group of 39-40, 41-42 and 43-44, respectively [36]. There is an increasing trend of the cost effectiveness from young age group to old age group.

With much larger sample size in this study, we revealed a significant decline trend of characteristics and treatment outcomes of IVF-ET with age increase. Data was stratified into three main age groups of <35 years old, 35-41 years old and ≥42 years old. Large sample size provided more power to our analysis. Number of previous cycles and duration of infertility increased with age increase. Basic FSH increased to larger than 10 IU/I when age ≥42, which indicated a decline of ovarian reserve after 42 years old. The same circumstance happened in basic AFC and AMH value. When age larger than 42 years, the IVF cycle cancellation rate got as large as 85.03%, which shows that only less than 15% cycles for women older than 42 got into embryo transfer procedure. And for 35-41 age group, the rate is about 50%. Also, with number of oocytes retrieved decreased by nearly half from <35 to 35-42, from 35-42 to ≥42 (p<0.05), the number of fertilized oocytes would be affected by the previous variable. Further stratification was conducted to data older than 42 years old. Basic FSH of 42, 43, 44 and ≥45 years old groups were all larger than 10, and 43 and ≥45 years groups showed higher values. Basic AFC reduced to about 3 after age of 44. And the cancellation rate was as high as 91.15% when age larger than 44. Number of oocytes retrieved decreased with age increases (p<0.05).

The quantity and quality decrease of oocytes lead to declined performance of IVF treatment. Our analysis showed that the number of pregnancy cycles decreased extremely after age 41 (from 5,673 to less than 200) and even less after age 45 (15 cycles in age 47). Biochemical pregnancy rate decreased since the basic number of cycles decreasing trend was large. Clinical pregnancy, live birth rate and multiple pregnancy rate all decreased rapidly after 42 years and the treatment outcome was worse after 45 years. Specifically, clinical pregnancy rate was 17.65% in 42 years old group, decreased almost half of clinical pregnancy rate in 35-41 years old group (37.85%). And this value decreased into 5.13% in 45 years old group. Live birth rate decreased over 70% from age 35-41 to 42 (24.47% to 7.06%). And this value decreased to 0 in 45 years old group, which means although there were 2 cycles got successful clinical pregnancy, the patients did not achieve live birth. For the 1 cycle in 47 years old group which got successful live birth, since the sample size is too small, this observation cannot stand for general statistical conclusion. From the analysis of this study, we can see that if take 42 years old as advanced maternal age, the characteristics and treatment outcome of IVF-ET would get worse with age increase. As for the economic cost analysis in our study, we only collected the roughly cost of all cycles and estimated the cost per live birth and get the approximate costs of IVF patients in single IVF center. The costs were 34,375.94 RMB for age group of <35 and increased nearly 6 times in age group of 42. When age larger than 44, the cost increased to 1357,154.07 RMB per live birth and this is a very large amount too average families in China. Our conclusion shows that IVF cost would increase when patient age increases.

There are three limitations in this study. One is the retrospective design of the study. The second limitation is that the period of the data collection ranged about 10 years, and in this period the ART technology has developed very fast and the inconsistency of variables may introduce bias into the data. The third limitation is within the economic cost data collection. These data were analyzed without considering the different subgroups of patients such as patients in different COS strategies. The costs for medicine in different treatment strategies are with large amount of difference. If we want to compare cost effectiveness of IVF in China to other countries, more information of IVF-ET treatment should be collected and included in the analysis.

## Conclusion

This study provided important conclusions for the relationship of maternal age with clinical characteristics and IVF-ET treatment outcomes. It can be helpful for the consultation in IVF centers. For female patients older than 42 years old especially 45 years old, they might intend to get unsatisfied outcome and higher cost per child. With the global trend of older age pregnancy and infertility treatment, more guidance and considerations should be focused on encouraging earlier age diagnosis and treatment.

## Data Availability

The datasets used and/or analyzed during the current study are available from the corresponding author on reasonable request.

## List of abbreviations

IVF-ET: In Vitro Fertilization and Embryo Transfer
ART: Assisted Reproductive Technology
AMA: Advanced Maternal Age
BMI: Body Mass Index
HCG: Human Chorionic Gonadotropin
FSH: Follicle Stimulating Hormone
AFC: Antral Follicle Count
AMH: Anti-Müllerian Hormone
COS: Controlled Ovarian Stimulation

## Ethics approval

The ethics committee of Jinjiang District Maternal and Child Health Hospital IVF center in Chengdu, China. has reviewed and approved this research.

## Consent for publication

“Not applicable” in this section.

## Authors’ contributions

ZL conducted data analysis, manuscript writing and results interpretation; ST conducted study design and part of data analysis; SL contributed to study design and part of analysis; HX provided clinical and laboratory evidence to the manuscript and part of data collection; ZL conducted data extraction; YZ conceived the study, manuscript writing and is the corresponding author of this manuscript. All the authors read and approved the final manuscript.

## Reference

[1] Barati E, Nikzad H, Karimian M. Oxidative stress and male infertility: Current knowledge of pathophysiology and role of antioxidant therapy in disease management. Cellular and Molecular Life Sciences, 2020: 1–21.

[2] World Health Organization. Reproductive health indicators for global monitoring: report of the second interagency meeting, WHO Geneva, 17-19 July 2000. World Health Organization, 2001.

[3] Zhou Z, Zheng D, Wu H, Li R, Xu S, Kang Y, et al. Epidemiology of infertility in China: a population□based study. BJOG: An International Journal of Obstetrics & Gynaecology, 2018, 125(4): 432–441.

[4] Crawford GE, Ledger WL. In vitro fertilisation/intracytoplasmic sperm injection beyond 2020. BJOG: An International Journal of Obstetrics & Gynaecology, 2019, 126(2): 237–243.

[5] Evenson D P, Djira G, Kasperson K, Christianson J. Relationships between the age of 25,445 men attending infertility clinics and sperm chromatin structure assay (SCSA®) defined sperm DNA and chromatin integrity. Fertility and Sterility, 2020, 114(2): 311–320.

[6] Sauer MV. Reproduction at an advanced maternal age and maternal health. Fertility and sterility, 2015, 103(5): 1136–1143.

[7] Ubaldi FM, Cimadomo D, Vaiarelli A, Fabozzi G, Venturella R, Maggiulli R, et al. Advanced maternal age in IVF: still a challenge? The present and the future of its treatment. Frontiers in endocrinology, 2019, 10: 94.

[8] Gleicher N, Kushnir VA, Albertini DF, Barad DH. Improvements in IVF in women of advanced age. Journal of Endocrinology, 2016, 230(1): F1–F6.

[9] Franasiak JM, Forman EJ, Hong KH, Werner MD, Upham KM, Treff NR, et al. The nature of aneuploidy with increasing age of the female partner: a review of 15,169 consecutive trophectoderm biopsies evaluated with comprehensive chromosomal screening. Fertility and sterility, 2014, 101(3): 656-663. e1.

[10] Capalbo A, Hoffmann ER, Cimadomo D, Maria Ubaldi F, Rienzi L. Human female meiosis revised: new insights into the mechanisms of chromosome segregation and aneuploidies from advanced genomics and time-lapse imaging. Human Reproduction Update, 2017, 23(6): 706–722.

[11] Martin JA, Hamilton BE, Sutton PD, Ventura SJ, Menacker F, Munson ML. Births: final data for 2002. National vital statistics reports, 2003, 52(10): 1–113.

[12] Mills M, Rindfuss RR, McDonald P, Te Velde E. Why do people postpone parenthood? Reasons and social policy incentives. Human reproduction update, 2011, 17(6): 848–860.

[13] Schmidt L, Sobotka T, Bentzen JG, Nyboe Andersen A, ESHRE Reproduction and Society Task Force. Demographic and medical consequences of the postponement of parenthood. Human reproduction update, 2012, 18(1): 29–43.

[14] Gleicher N, Kushnir VA, Weghofer A, Barad DH. The “graying” of infertility services: an impending revolution nobody is ready for. Reproductive Biology and Endocrinology, 2014, 12(1): 63.

[15] Adamson GD, de Mouzon J, Chambers GM, Zegers-Hochschild F, Mansour R, Ishihara O, et al. International Committee for Monitoring Assisted Reproductive Technology: world report on assisted reproductive technology, 2011. Fertility and sterility, 2018, 110(6): 1067–1080.

[16] Li YH, Wang YP, Dai L, Zhou GX, Liang J, Li Q, et al. The trend of national advanced maternal age woman proportion in hospital-based surveillance. Zhonghua yu Fang yi xue za zhi [Chinese Journal of Preventive Medicine], 2009, 43(12): 1073–1076.

[17] Jiang L, Chen Y, Wang Q, Wang X, Luo X, Chen J, et al. A Chinese practice guideline of the assisted reproductive technology strategies for women with advanced age. Journal of Evidence□Based Medicine, 2019, 12(2): 167–184.

[18] Reichman D, Rosenwaks Z. GnRH antagonist-based protocols for in vitro fertilization. In: Human Fertility. Humana Press, New York, NY, 2014: 289–304.

[19] Huang J Y J, Rosenwaks Z. Assisted reproductive techniques. In: Human Fertility. Humana Press, New York, NY, 2014: 171–231.

[20] Annan JJK, Gudi A, Bhide P, Shah A, Homburg R. Biochemical pregnancy during assisted conception: a little bit pregnant. Journal of clinical medicine research, 2013, 5(4): 269.

[21] Gunnala V, Irani M, Melnick A, Rosenwaks Z, Spandorfer S. One thousand seventy-eight autologous IVF cycles in women 45 years and older: the largest single-center cohort to date. Journal of Assisted Reproduction and Genetics, 2018, 35(3): 435–440.

[22] Ramirez LB, Klein JU, França UL. Assisted reproduction care concentration trends in the united states. Fertility and Sterility, 2018, 110(4): e269.

[23] https://www.cdc.gov/obesity/adult/defining.html

[24] GleicherN, Kushnir VA, Albertini DF, Barad DH. Improvements in IVF in women of advanced age. Journal of Endocrinology, 2016, 230(1): F1–F6.

[25] Practice Committee of the American Society for Reproductive Medicine. Aging and infertility in women. Fertility and sterility, 2006, 86(5 Suppl 1): S248.

[26] Maheshwari A, Hamilton M, Bhattacharya S. Effect of female age on the diagnostic categories of infertility. Human reproduction, 2008, 23(3): 538–542.

[27] Hsieh HC, Su JY, Wang S, Huang YT. Age effect on in vitro fertilization pregnancy mediated by anti-Mullerian hormone (AMH) and modified by follicle stimulating hormone (FSH). BMC pregnancy and childbirth, 2020, 20: 1–8.

[28] Broekmans FJ, Kwee J, Hendriks DJ, Mol BW., Lambalk CB. A systematic review of tests predicting ovarian reserve and IVF outcome. Human reproduction update, 2006, 12(6): 685–718.

[29] Wiweko B, Prawesti DMP, Hestiantoro A, Sumapraja K, Natadisastra M, Baziad A. Chronological age vs biological age: an age-related normogram for antral follicle count, FSH and anti-Mullerian hormone. Journal of assisted reproduction and genetics, 2013, 30(12): 1563–1567.

[30] Barbakadze L, Kristesashvili J, Khonelidze N, Tsagareishvili G. The correlations of anti-mullerian hormone, follicle-stimulating hormone and antral follicle count in different age

[31] Van Rooij Iaj, Broekmans Fjm, Te Velde ER, Fauser Bcjm, Bancsi Lfjmm, De Jong FH, Themmen APN. Serum anti-Müllerian hormone levels: a novel measure of ovarian reserve. Human Reproduction, 2002, 17(12): 3065–3071.

[32] Barad DH, Weghofer A, Gleicher N. Comparing anti-Müllerian hormone (AMH) and follicle-stimulating hormone (FSH) as predictors of ovarian function. Fertility and sterility, 2009, 91(4): 1553–1555.

[33] Szamatowicz M, Grochowski D. Fertility and infertility in aging women. Gynecological Endocrinology, 1998, 12(6): 407–413.

[34] Stern JE, Cedars MI, Jain T, Klein NA, Beaird CM, Grainger DA, Gibbons WE. Assisted reproductive technology practice patterns and the impact of embryo transfer guidelines in the United States. Fertility and sterility, 2007, 88(2): 275–282.

[35] Klipstein S, Regan M, Ryley DA, Goldman MB, Alper MM, Reindollar RH. One last chance for pregnancy: a review of 2,705 in vitro fertilization cycles initiated in women age 40 years and above. Fertility and sterility, 2005, 84(2): 435–445.

[36] Audibert C, Glass D. A global perspective on assisted reproductive technology fertility treatment: an 8-country fertility specialist survey. Reproductive Biology and Endocrinology, 2015, 13(1): 1–13.

